# Rare Diseases Clinical Trials Toolbox - Public resources and main considerations to set up a clinical trial on medicinal products for humans in Europe

**DOI:** 10.1101/2024.02.15.24302854

**Authors:** Marta del Álamo, Biljana Zafirova, Martina Esdaile, Sarah Karam, Sabine Klager, Christine Kubiak

**Affiliations:** ECRIN, European Clinical Research Network Infrastructure, Paris, France

**Keywords:** Rare diseases, clinical trial, paediatric, toolbox, resources, academic-sponsored, investigator-initiated

## Abstract

**Background:** Drug development programmes in rare diseases have many challenges, some of which differ from those facing researchers working on common diseases, like the scarcity of patients.

Over the past years, research and regulatory initiatives, as well as resources have been implemented to expedite drug development for rare diseases. Nevertheless, these tools have been developed in the context of different projects and with diverse aims. Therefore, they have not yet been structured to encompass the conduct of clinical trials as a whole. To address this issue, the EJP RD (European Joint Program for Rare Diseases) has developed the Rare Diseases Clinical Trial Toolbox.

**Purpose:** This toolbox collates the accumulated knowledge, experience, and resources (collectively termed ‘tools’) generated by projects, research infrastructures and/or other organizations into a structured, practical and guided instrument to help clinical trialists and trial managers understand the regulations and requirements for conducting trials, with a special focus on investigator-initiated trials for rare diseases.

**Methods:** The toolbox is organized into five domains: research question, plan, execution, analysis, and end of trial. Each domain describes one or several activities to be considered in this step and indicate at what stage of the trial pathway these activities should take place, regardless of the therapeutic area. Each activity is further linked to specific resources (the tools) that are relevant for those activities. Associated resources are in the public domain developed in the context of research projects or by relevant clinical research stakeholders. Selected tools must be of fundamental importance to clinical trials and be applicable to *rare diseases clinical research*. Rare-diseases specific resources are highlighted as such and include those specially relevant to paediatric clinical research, considering than one half of rare disorders affect children and some 60% of designated orphan medicines are intended for paediatric use.

**Results:** The current version of the Toolbox includes 111 resources tagged as relevant for any of the 18 activities within the clinical trial outline. Overall, 75 % of all resources are relevant to any clinical trial while 25 % are tagged as “rare disease specific”.

**Conclusion:** Access to public resources relevant to the development of clinical trials for rare diseases is sometimes challenged by limited awareness and/or the absence of an adequate framework that enables their findability. This Toolbox aims at building a framework supporting the optimal use of existing tools.

## Background

Drug development programmes in rare diseases (RD) have many challenges, some of which differ from those facing researchers working on common diseases, such as the lack of clinical research experts and the scarcity of patients (1–3). In the RD context, multinational, multi-centre clinical trials are required to achieve sufficient recruitment and provide added value by promoting global standards of care and increasing the applicability of the research findings. International collaboration is nevertheless constrained by scientific, ethical, financial and regulatory considerations. Different sponsors may have varying capacities to address these constraints, leading to divers collaborative patterns.

Commercially sponsored clinical trials are at the origin of bringing most of the new drugs to market. However, these clinical trials only assess the safety and efficacy of drugs that are chosen by the commercial entity which funds the entire process. Non-commercial or academic investigator-initiated trials therefore have their own specific significance, often focusing on new or refining indications of available treatments and optimizing therapeutic strategies that do not contribute to significant financial gain to the pharmaceutical industry.

In contrast to industry-sponsored trials, academic-sponsored trials face a number of challenges, including: (a) lack of funding, or flexible, long-term financial support covering unexpected issues that arise during the trial (b) inadequate infrastructure to plan and execute the trial including a working quality management system (c) insufficiently structured processes to facilitate academic collaborations, and a lack of platforms to discuss and solve issues related to academic trials that result in difficulties accessing the right partner assisting on the operational management (usually provided by contract research organizations (CROs) for industry-sponsored trials).

Academic sponsors and investigators end up involved, not only in the scientific aspects of the research, but also in navigating the operational coordination and management as well.

With this Rare Diseases Clinical Trials Toolbox (4) we aim to provide a guide for researchers, charities and other academic trials stakeholders embarking on the set up of clinical trials for rare diseases. We highlight common activities, issues and considerations at each development stage and signpost to available tools.

## Methods

The RD Clinical Trials toolbox aims to collates the accumulated knowledge, experience, and resources (collectively termed as ‘tools’) developed in the context of previous research projects and/or by research infrastructures and other organizations into a practical and guided toolbox to help clinical trialists and clinical research managers understand the regulations and requirements for conducting trials, with special focus on investigator-initiated trials for rare diseases. Rare-diseases specific resources include those specially relevant to paediatric clinical research, considering than one half of rare disorders affect children and some 60% of designated orphan medicines are intended for paediatric use.

The Toolbox does not aim to be a database of existing resources, but to classify and structure them in the context of the different processes and activities to be considered while setting up a clinical trial. Thus, five steps were followed to develop the Toolbox (Fig. 1)

**Fig. 1.**
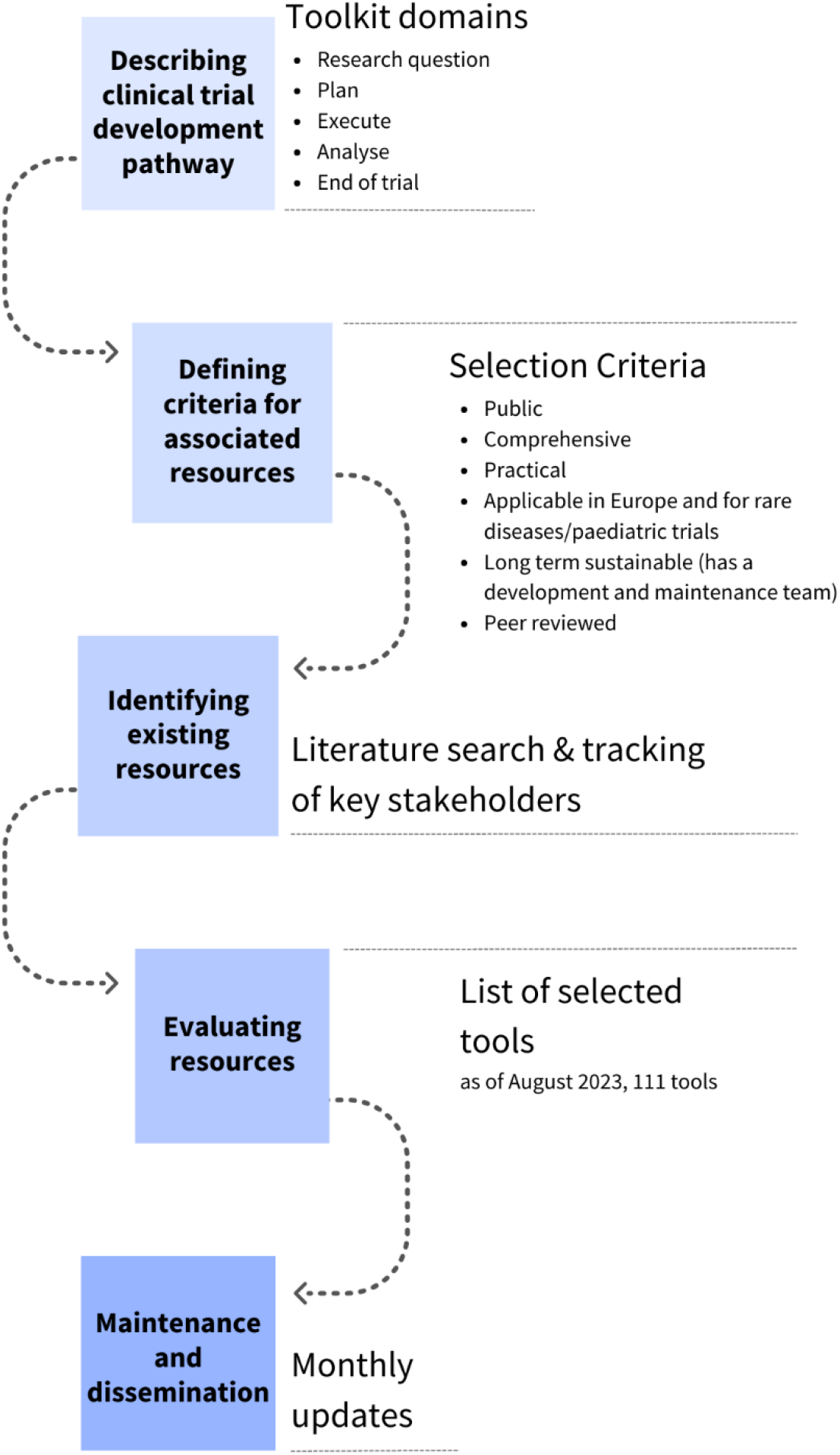
Development and implementation of the RD Clinical Trials Toolbox.

### Step 1-Describing the Clinical Trial Development Pathway

The clinical trial pathway is not specific to rare disease and is typically defined by five main domains. These same domains are used to structure the Toolbox:

- Research question
- Planning
- Execution
- Analysis
- End of the trial

Each domain describes one or several activities that occur through the trial pathway and that need to be considered, regardless of the therapeutic area (Figure 2).

**Fig 2.**
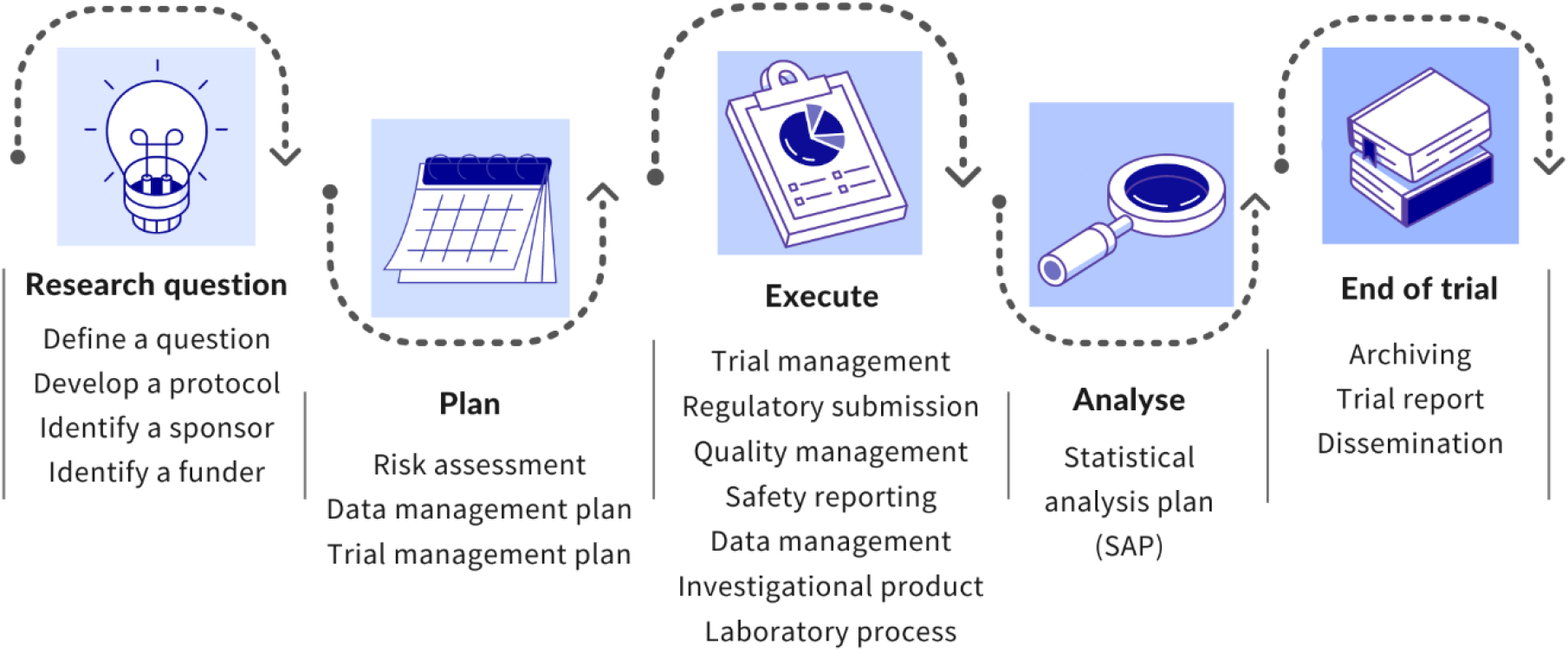
RD Clinical Trials Toolbox outline.

For each activity a detailed description of their relevance and any necessary considerations are provided. The relevance of the selected activities for any clinical trial and any rare disease-specificity is described based on the scope of the toolbox.

Scope: This Toolbox has been developed to cover geographically the European Union (as per applicable regulation) and to focus on medicinal products for human use. Although it is acknowledged that medical devices and advanced therapy products are especially relevant for rare diseases research, the regulatory framework of clinical studies/trials for these investigational products is still complex and not fully harmonized in Europe and beyond. The recent implementation of the Clinical Trials Regulation (5) in Europe works to harmonize the clinical trial regulatory landscape in Europe for medicinal products. Thus, we have considered that the selected scope of medicinal products for human use was best suited for a first version of the Toolbox, which is planned to cover other interventions (ATMP, medical devices) in the future. Within the clinical trials pathway, the breadth of the toolbox extends from the initial trial design to final trial results reporting and dissemination.

### Step 2-Defining the criteria for associated resources

Each domain and activity is further elaborated through the identification of resources that are relevant for the specific step. Associated resources are in the public domain: documents, guidance notes,, templates, and other research outcomes developed in the context of previous research projects, and/or developments of relevant clinical research stakeholders as research infrastructures, universities, patient organizations. Selected tools, standards, and guidelines must be of fundamental importance to clinical trials and with special focus on rare diseases clinical research. Tools which are non-rare-diseases specific are eligible if still relevant to the setup of clinical trials for rare diseases. Final selection criteria to determine the relevance of tools, standards, and guidelines for the Toolbox, are:

- Public
- Comprehensive
- Practical
- Applicable in Europe and for rare diseases trials
- Long term sustainability (has a development and maintenance team)
- Peer reviewed

(National, regional or institutional resources for rare diseases, or a specific single disease entity, or commercial resources are excluded from the selection, although acknowledged as being also very important.

### Step 3-Identifying Tools

A comprehensive, yet not systematic, search in English was performed to find potential tools that have been developed within publicly funded projects and/or by research infrastructures and organizations dedicated to clinical research, using the toolbox domains as keywords. Main sources of information include, in addition to ICH and EMA guidelines, current and past research programs for rare diseases (including paediatrics) (IRDiRC, EJP RD, PedCRIN), European research infrastructures (ESFRI roadmap) and patient engagement organizations (EUPATI, EURORDIS). Tools are matched against the above-mentioned selection criteria.

### Step 4-Evaluating Tools

Pre-identified resources fulfilling the selection criteria are categorized under the relevant domain and activity of the clinical trial pathway. Relevance of each resource and rare-diseases specificity is described. Selected resources are accessible through their original location using public links. Tools developed within EU funded projects are accessible long-term through the EU CORDIS website (6).

### Step 5-Maintenance and dissemination

The Toolbox is a living tool, hence new or currently unidentified resources that come to the light will be added or modified as required. The Toolbox is hosted on the ECRIN website (4). It has been labelled as an IRDiRC (International Rare Diseases Research Consortium) recognized resource and is accessible through IRDiRC website (7).

## Results

The current version of the toolbox includes outlines of the 18 activities and 111 tools (https://ecrin.org/sites/default/files/inline-files/Toolbox_August%202023%20BZ.pdf) tagged as relevant for any of the activities within the clinical trial outline.

The domain “execute” contains the most tools, 50 % (56), followed by domain “research question” with 34 tools (Fig. 3).

**Fig 3.**
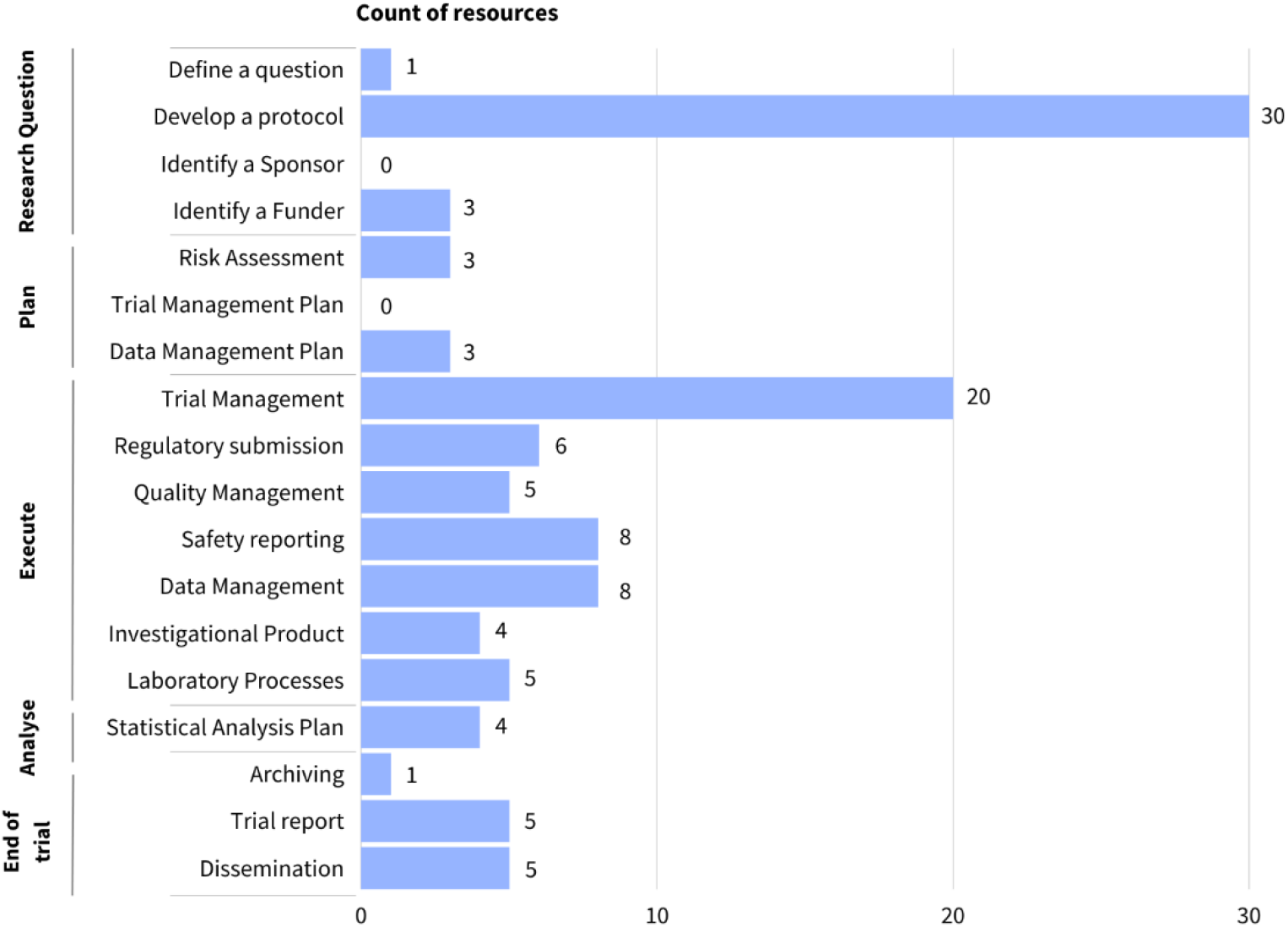
Number of resources in Toolbox per domain and activity at key steps of the clinical trial development pathway.

Among the resources addressing activities linked to the “execution” of the trial, 14 are rare-disease specific, 20 are on trial management, eight are on data management and 8 cover on safety reporting.

In the “research question” domain, the most represented activity is “develop a protocol” with 30 tools, of which 16 are rare-disease specific.

The Toolbox includes a variety of different types of resources. Thirty-eight percent or 42 resources are guidelines, followed by 10% or 11 resources that are recommendations and equal of toolboxes (Fig. 4)

**Fig 4.**
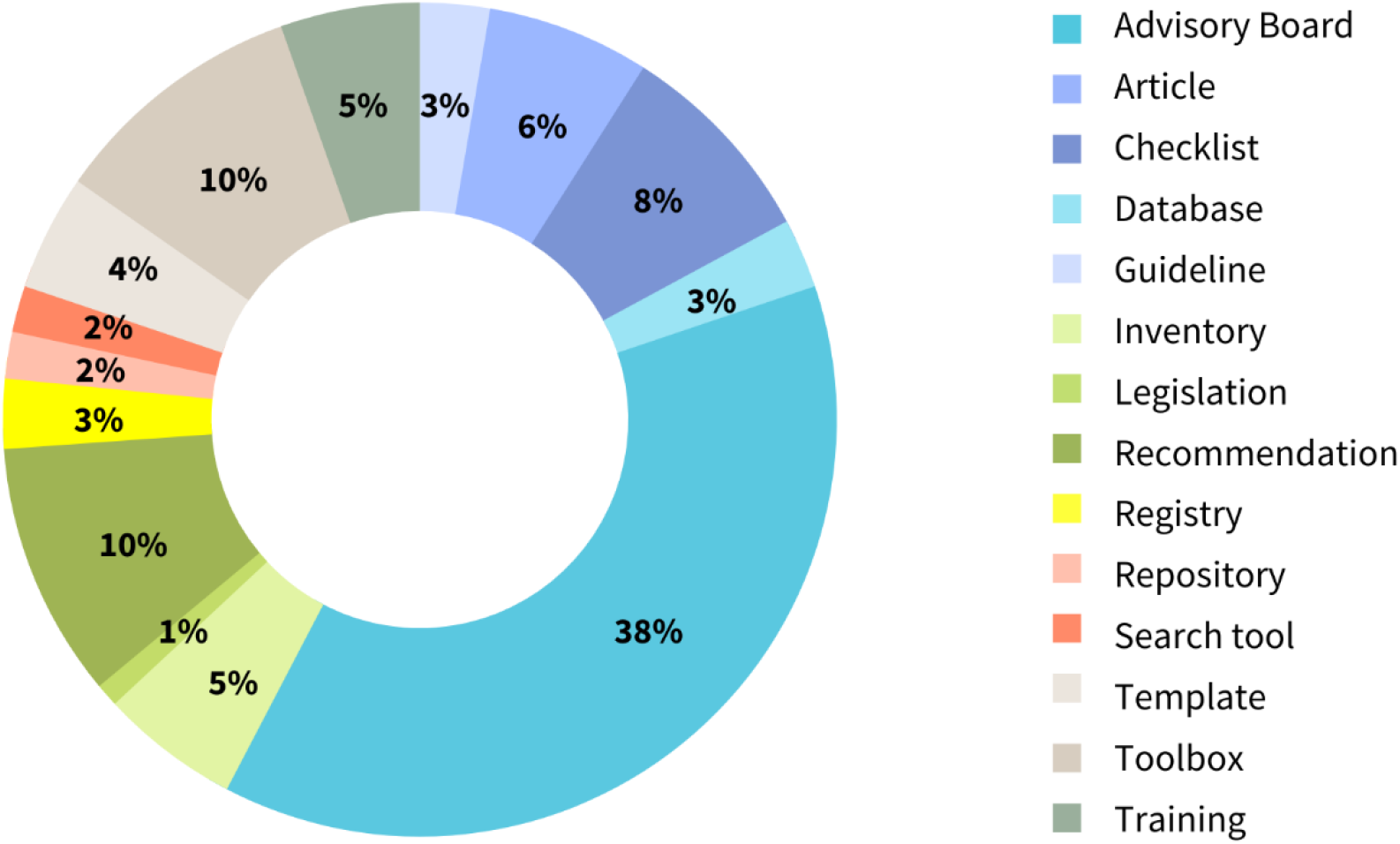
Representation of the different type of resources in the RD Clinical Trials Toolbox.

Overall, 72% of all resources are relevant to any clinical trial within the scope of the toolbox while 28% are tagged as “rare disease-specific”. From the latter, 14 resources are guidelines and five resources are recommendations that have been developed to fine-tune more general guidelines considering specificities of paediatrics trials (pharmacokinetics and ethics) and rare diseases (small number/patients’ heterogeneity).

## Discussion

A recent analysis of six use cases (1) describes obstacles hindering the development of academic-sponsored clinical trials for rare diseases, highlighting the high number of hurdles related to operational management. Academic sponsors and investigators end up getting involved not only in the scientific aspects of the research, but also having to navigate regulatory aspects, operational coordination and management themselves. Better information about existing resources such as research infrastructures, clinical research programs, and counselling mechanisms is needed to support and guide clinicians and sponsors through the many challenges associated to the set-up of academic-sponsored multinational trials. Over the past years, research, policy, regulatory initiatives and resources have been introduced to expedite drug development for rare diseases (2). Nevertheless, these tools have been developed in the framework of different projects and with different aims and therefore they have not yet been collated and structured to support the conduct of a clinical trials as a whole. To address this issue, the EJP RD (European Joint Program for Rare Diseases) has developed the Rare Diseases Clinical Trial Toolbox. This toolbox aims at building a framework in which existing tools applicable to the development of clinical trials for rare diseases can be presented.

### Existing toolboxes

Practical advice to researchers in designing and conducting trials has been previously published as guidelines, toolboxes, guidebooks etc. Addressing different goals, they differ mostly on their scope and format.

In the USA, the NIH maintains a clinical research study investigator’s toolbox (15), a Web-based informational repository for investigators and staff involved in clinical research. This toolbox contains templates, sample forms, guidelines, regulations, and informational materials to assist investigators in the development and conduct of high-quality clinical research studies in the USA. It applies a very practical approach, with a focus on providing examples of procedures and templates for general clinical research, without a specific outline for clinical trials.

In Europe, the NIHR Clinical Trials Toolbox was developed in 2003 by the UK Medical Research Council (MRC) and the UK Department of Health (16) as a tool to guide investigators embarking on a clinical trial through the regulatory and governance requirements. The NIHR Clinical Trials Toolbox is a good starting point for trialists and trial managers to ensure all legal obligations are met and presents the information as a structured Roadmap, but with a strong focus in the UK Clinical Trials Regulation and national specificities.

At international level, but with a clear focus on a specific disease, the Malaria Clinical Trials Toolbox (17) is presented as a pathway with a step-by-step guide for researchers on how to plan, design, execute and interpret malaria clinical trial results.

In the field of Rare Diseases, IRDiRC has developed and published, the Orphan Drug Development Guidebook (ODDG) (7), a patient focused guidebook that describes the available tools, incentives, resources and practices for developing traditional and innovative drugs/therapies for rare diseases and how to best use them. Structured as factsheets, the ODDG guides users through the full orphan drug development process, including regulatory pathways, HTA (Heath Technology Assessment) and reimbursement, early access and development practices and resources at international level.

With a different focus, the Rare Diseases Clinical Trial Toolbox integrates some of the tools already included in these previous initiatives and incorporates them, together with other resources, in a guided, practical structure as defined by the Clinical Trial outline.

The current version of the Rare Diseases Clinical Trial Toolbox is primarily focused on Clinical Trials of Investigational Medicinal Products (CTIMPs) in the EU and the regulatory environment and requirements associated with these.

### Generic clinical trials tools

Although rare diseases may present unique clinical problems, some of the methodological and operational challenges to studying health outcomes are common for other diseases areas. Indeed, generic guidelines relevant for rare diseases clinical trial design include the ICH general guidelines (18) and the main SPIRIT statement (19,20). Fine-tuned versions for paediatrics and/or rare diseases guidelines have subsequently been published as a complement to these specific guidelines. Other relevant resources, such as a clinical trials registry/metadata repository, are still missing their “rare-diseases”-specific version. Thus, by presenting all relevant tools in a structured framework, the Toolbox contributes to avoiding duplication and to uncovering possible gaps.

### Rare disease specific tools

Previous works have highlighted the need of specific methodologies for enabling meaningful data generation from trials in small populations and designing outcome measures that are focused on and relevant for patients (3,21). Indeed, several resources aiming to fill this gap include outcomes of the European funded projects IDEAL, ASTERIX and INSPIRE (22–24) regarding novel trial designs or the Patient Reported Outcome Measures (PROM) repository developed by the ERICA project. Other interesting initiatives aiming to provide support for rare diseases/paediatrics populations include advisory boards for patient involvement (EURORDIS) and Informed Consent Guidelines. Examples of the latter are: a) the model consent clauses developed by GA4GH and IRDiRC (25) for rare diseases research. It aims to improve data interoperability, to meet the informational needs of participants, and to ensure proper ethical and legal use of data sources and participants’ overall protection; b) the PedCRIN tool, Neonatal trials and informed consent (11), which provides a checklist of practical points to consider when talking to parents about the possible inclusion of a neonate into a clinical trial; c) the Assent/Informed Consent Guidance for Paediatric Clinical Trials with Medicinal Products in Europe developed by Enpr-EMA’s Working Group on Ethics. This document is intended to be used as an overview tool of the contents for assent/informed consent forms for all stakeholders (such as patients, sponsors and investigators) to support the conduct of high quality paediatric clinical trials in Europe across all paediatric age groups, from birth to less than 18 years of age (26).

### Activities with limited tools

As noted earlier, it is also essential to understand where there are gaps in existing support. Two activities within the “Research Question” domain have none (Identify a sponsor) or limited (Identify a funder) identified resources.

### Identify a funder

While this may be a less pertinent problem for industry-funded trials, funding has been identified as a main challenge for academic clinical trials. In the rare diseases area, the small patient population can dampen commercial interest, making the need to address this challenge all the more relevant. Indeed, public funding calls for multinational clinical trials might be scarce, difficult to identify and/or not suitable to support specific research questions.

Funders’ communication channels aim to reach investigators and trialist, but often funding is mostly restricted to national level, making difficult to identifying opportunities for the funding or development of multinational trials. A comprehensive database on clinical trial funders in Europe is missing. Interestingly, EU co-funded programs like ERA4Health (27) are currently working on building capacity in conducting investigator-initiated clinical studies, with plans to map the funding landscape in Europe.

The current version of the RD Clinical Trial Toolbox includes two tools on the topic. Both NIRO (Navigating Innovation & Research Opportunities) (28) and research funding database ScientifyResearch (29) have developed open, curated and structured research funding database to help researchers find funding opportunities. Although the scope of both tools is general research (with NIRO more focused on infectious diseases), their search engines allow narrowing the search at clinical trial level.

### Identify a sponsor

For non-industry sponsored trials, the sponsor is usually the institution to which the investigator is affiliated, a research foundation, and/or patient organization. A selection of these academic/public institutions have previous experience and specific personnel to work as sponsor’s representatives, but in other cases, the coordinating investigator assumes the sponsor’s role. To our knowledge, no database compiling information about academic sponsors exists and, unlike industry sponsors, academic institutions acting as sponsors are rarely present in European forums discussing clinical trials implementation. Interestingly, the current EU clinical trials transformation initiative: Accelerating Clinical Trials in the EU (ACT EU) (30) workplan aims to establish a process to support academic sponsors to undertake large multinational clinical trials.

## Conclusions

Academia plays a key role in the development of drugs for rare diseases (31) but sponsors and investigators leading these trials face a wide range of challenges that hinder patient’s access to drugs. Over the past years, public funding has supported initiatives, projects, and organizations to make gap analysis (1,32,33) and to develop resources supporting and guiding clinicians through the many challenges associated to the set-up of academic-sponsored trials. Access to these public resources is sometimes challenged by limited awareness and/or absence of an adequate framework that make them findable. The RD Clinical Trial Toolbox is geared to support this community and provide a framework in which they can navigate the various tools that have been made available.

## Data Availability

All data produced are available online at https://ecrin.org/rare-diseases-clinical-trials-toolbox

https://irdirc.org/resources-2/irdirc-recognized-resources/

https://imt.ejprarediseases.org/collection/clinical-trials-toolkit/

## List of abbreviations

ACT EU: Accelerating Clinical Trials in the EU

CTIMPs: Clinical Trials of Investigational Medicinal Products

CRO: contract research organizations

IRDiRC: International Rare Diseases Research Consortium

NIRO: Navigating Innovation & Research Opportunities

ICH: International Council for Harmonization

EMA: European Medicines Agency

EJP: European Joint Program for Rare Diseases

ESFRI: Strategy Forum on Research Infrastructures

EUPATI: European Patients Academy on Therapeutic Innovation

EURORDIS: European Organization for Rare Diseases

ODDG: Orphan Drug Development Guidebook

PedCRIN: Paediatric Clinical Research Infrastructure Network

PROM: Patient Reported Outcome Measures

RD: Rare diseases

## Declarations

### Availability of data and materials

All data supporting the findings of this study are available within the paper and the following websites:

https://ecrin.org/rare-diseases-clinical-trials-toolbox

https://www.ejprarediseases.org/rare-diseases-clinical-trials-toolbox/

https://imt.ejprarediseases.org/collection/clinical-trials-toolkit/

A list of all resources can be downloaded through all the above mentioned websites

### Ethics approval and consent to participate

Not applicable.

### Consent for publication

Not applicable.

### Competing interests

The authors declare that they have no competing interests

### Funding

ECRIN has received funding from the European Union’s Horizon 2020 research and innovation program under the EJP RD COFUND-EJP N° 825575 to develop the Rare Diseases Clinical Trials Toolbox.

## Acknowledgments

This article was written on behalf of the contributors to the Rare Diseases Clinical Trials Toolbox. The contributors beyond the authors were Ralf-Dieter Hilgers, Rima Nabbout and Agustin Arasanz Duque.

## Authors’ contribution

MdA, SKL, CK and ME participated in the design and development of the toolbox. MdA, ME and BZ search, curate and update the tools. MdA and BZ drafted the initial version of the manuscript. ME and SKA created the figures. All authors participated in the discussions, read and critically revised the manuscript for important intellectual content, and approved the final version for publication.

## Notes

### Competing Interest Statement

The authors have declared no competing interest.

### Funding Statement

ECRIN has received funding from the European Union Horizon 2020 research and innovation program under the EJP RD COFUND-EJP N 825575 to develop the Rare Diseases Clinical Trials Toolbox

